# Shared Pathogenic Pathways Between REM Sleep Behavior Disorder and Neurodegenerative and Psychiatric Disorders

**DOI:** 10.64898/2026.02.05.26345599

**Authors:** Zhao Zhang, Emma N. Somerville, Zih-Hua Fang, Lang Liu, Farnaz Asayesh, Jamil Ahmad, Saeid Amiri, Meron Teferra, Pauline Dodet, Isabelle Arnulf, Michele T.M. Hu, Alex Desautels, Yves Dauvilliers, Merve Aktan-Süzgün, Abubaker Ibrahim, Ambra Stefani, Birgit Högl, Carles Gaig, Angelica Montini, Gerard Maya, Alex Iranzo, Monica Serradell, Gian Luigi Gigli, Mariarosaria Valente, Francesco Janes, Andrea Bernardini, Karel Sonka, David Kemlink, Petr Dusek, Michael Sommerauer, Sinah Röttgen, Michela Figorilli, Monica Puligheddu, Brit Mollenhauer, Claudia Trenkwalder, Friederike Sixel-Doring, Giuseppe Plazzi, Francesco Biscarini, Elena Antelmi, Valerie Cochen De Cock, Michele Terzaghi, Giuseppe Fiamingo, Anna Heidbreder, Luigi Ferini-Strambi, Miriam Ostrozovicova, Matej Skorvanek, Kristina Kulcsarova, Jitka Buskova, Beatriz Abril, Beatrice Orso, Pietro Mattioli, Dario Arnaldi, Bradley F. Boeve, Yo-El Ju, Owen A. Ross, Shih-Ying Wu, Jonghun Lee, Daria Prilutsky, Cornelis Blauwendraat, Hampton Leonard, Ronald B. Postuma, Guy A. Rouleau, Ziv Gan-Or

## Abstract

Isolated/idiopathic rapid-eye-movement (REM)-Sleep Behavior Disorder (iRBD) is characterized by dream enactment behaviors associated with loss of REM atonia. iRBD is in most cases a prodromal synucleinopathy, and emerging evidence suggests associations between RBD and other neurological and psychiatric conditions. In this study, we performed pathway-based polygenic risk score (PRS) and rare variant burden analyses to examine these potential associations. Pathway-specific PRS were constructed from genome-wide association study summary statistics of five neurodegenerative and seven psychiatric traits across 10 biologically relevant pathway categories, including a total of 279 pathways, in 1,573 iRBD cases and 16,022 controls from the International RBD Study Group and UK Biobank. Rare variant burden tests were performed in 1,264 iRBD cases and 2,581 controls. We identified multiple potential pathways indicating shared polygenic risk between RBD and both neurodegenerative and psychiatric disorders. Lewy body diseases and post-traumatic stress disorder had the most shared polygenic risk pathways in neurological and psychiatric disorders, respectively. Two pathways, the serotonin transport pathway and the chaperone-mediated autophagy pathway, showed the strongest association with iRBD, and gene-based rare variants analyses revealed five genes associated with iRBD: *GBA1*, *PLEKHM1*, *LRP2*, *P2RX1*, and *HAP1.* Subsequent analysis of these genes in Parkinson’s disease and dementia with Lewy bodies replicated several associations. Together, these findings provide novel insights into the shared genetic architecture underlying iRBD, neurodegenerative disorders, and psychiatric traits, with implications for early identification and mechanistic understanding.

## Introduction

Isolated/idiopathic rapid-eye-movement (REM) Sleep Behavior Disorder (iRBD) is characterized by loss of normal muscle atonia during REM sleep, leading to dream enactment behaviors^1^. RBD has attracted considerable attention due to its strong association with synuclein-related neurodegenerative disorders, including Parkinson’s disease (PD), dementia with Lewy bodies (DLB), and multiple system atrophy (MSA)^2, 3^. Longitudinal studies indicate that up to 73.5% of individuals with iRBD convert to parkinsonism or dementia within 12 years, with more converting after longer follow-up^4^.

Although RBD is more frequent in α-synucleinopathies, several reports suggest that a minority of patients may develop Alzheimer’s disease (AD)^5^, and RBD can occur in a small subset of amyotrophic lateral sclerosis (ALS) patients^6, 7^. RBD has also been found to be 10-fold more likely to develop in patients with psychiatric disorders^8^. While antidepressants have been implicated as potential triggers, affecting up to 6% of users, RBD can persist even after discontinuation of medications, suggesting an underlying, preexisting vulnerability^9^. Moreover, patients with confirmed iRBD exhibited a higher prevalence of early-onset psychiatric disorders, including depression, anxiety, and Post-Traumatic Stress Disorder (PTSD)^10^. Together with evidence for overlapping mechanisms between neurodegenerative and major psychiatric disorders^11^, these observations suggest that shared genetic or mechanistic factors may underlie RBD and psychiatric disease.

Here we test the hypothesis that RBD and neurodegenerative or psychiatric disorders have a shared genetic basis, and aim to identify shared biological pathways, using the largest genetic cohort of iRBD.

## Results

### Pathway-specific PRS identifies multiple potential pathways shared between RBD and other diseases

Pathway-specific PRS analysis identified 58 pathways derived from neurodegenerative or psychiatric disorders that were associated with iRBD after false discovery rate (FDR) correction on the raw p-values (Supplementary Table S1). Notably, eight pathways derived from Lewy body dementia (LBD) GWAS showed positive associations with RBD susceptibility and remained significant after stringent empirical FDR correction, representing the only pathways to meet this threshold (Fig. 1).

**Figure 1.**
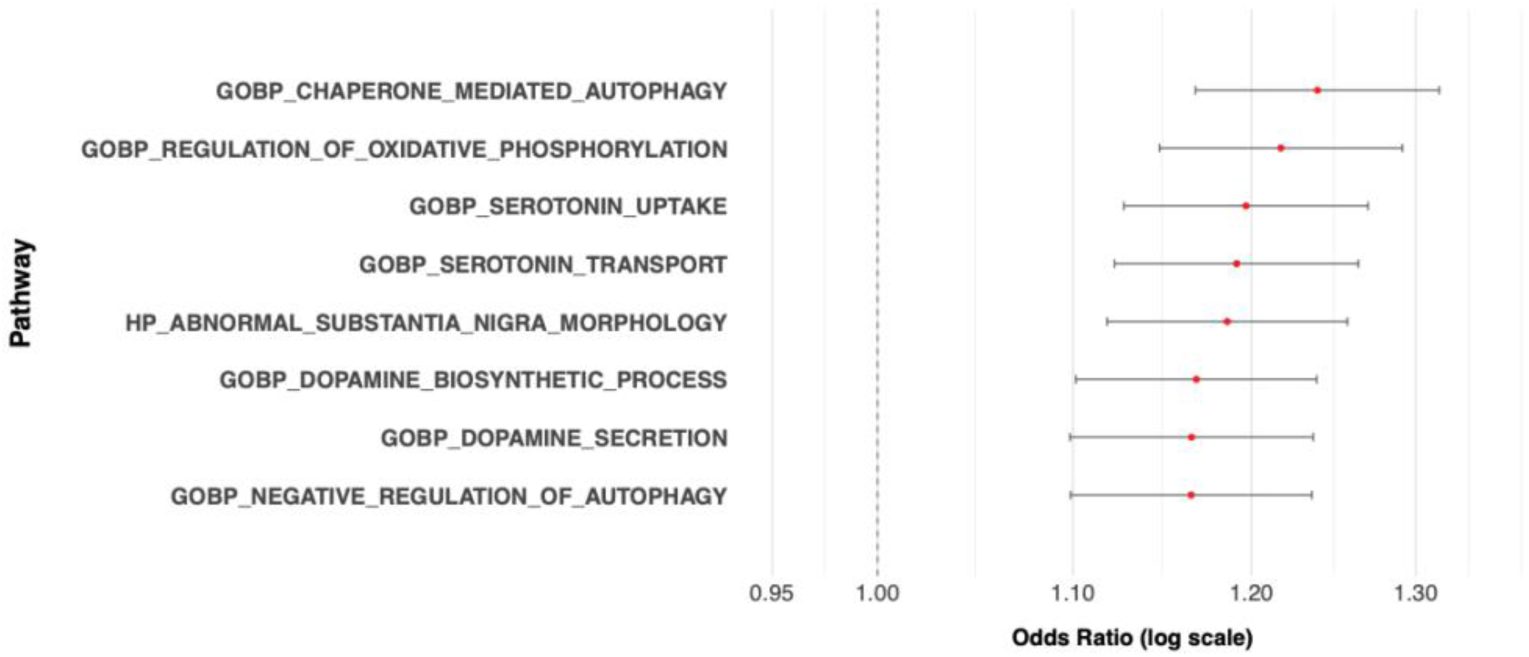
Forest plot of LBD-derived pathways positively associated with RBD susceptibility. Odds ratios (ORs) and 95% confidence intervals (CIs) were estimated from pathway-specific PRS. Eight pathways derived from LBD GWAS remained significant after empirical FDR correction.

Among the 58 significant pathways, those derived from neurodegenerative disorder GWAS were predominantly enriched for neurotransmission and synaptic function, whereas fewer pathways were associated with autophagy/lysosomal function, mitochondrial function, immune response, and calcium signaling (Fig.2, Supplementary Table S2.1). Pathways derived from LBD GWAS exhibited the most extensive enrichment across all disorders, including neurotransmission pathways (dopaminergic, glutamatergic, GABAergic, and acetylcholine signaling), mitochondrial oxidative phosphorylation, calcium signaling, autophagy/lysosomal function, and immune response.

**Figure 2.**
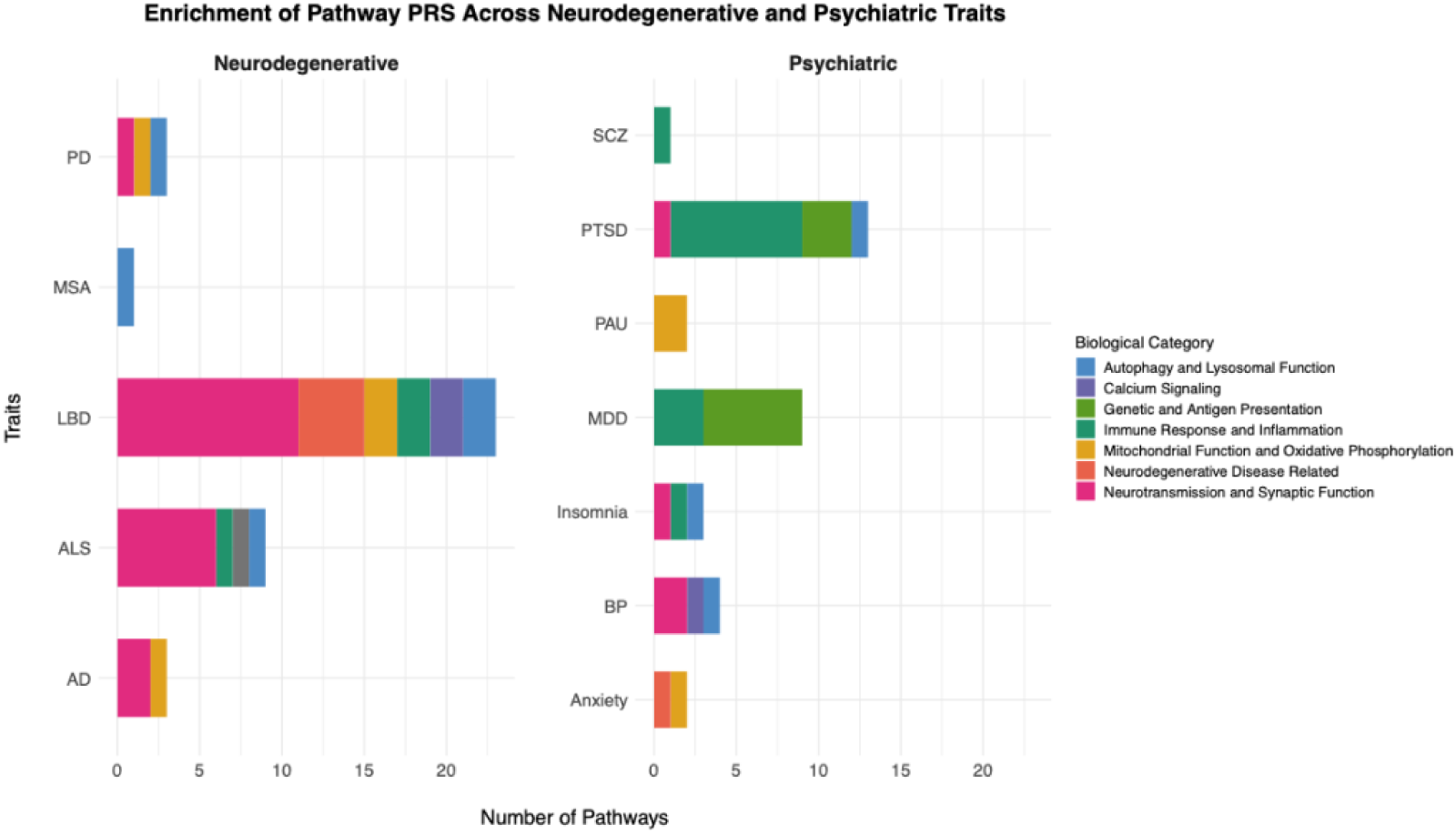
Enrichment of Pathway PRS Across Neurodegenerative and Psychiatric Traits. Among 58 significant pathways, those derived from neurodegenerative disorder GWAS were predominantly enriched for neurotransmission and synaptic function. Psychiatric trait pathways showed more heterogeneous enrichment.

Psychiatric traits displayed more heterogeneous enrichment profiles. Among them, pathways derived from PTSD GWAS showed the most extensive enrichment, primarily in immune response pathways, including B cell activation, lymphocyte activation, general immune effector functions, and adaptive immune system pathways. Additional contributions were observed from genetic and antigen presentation pathways, particularly MHC class II–mediated antigen processing, as well as autophagy/lysosomal pathways and neurotransmission via the acetylcholine release cycle (Supplementary Table S2.2). Other pathways derived from psychiatric disorders GWAS are detailed in Supplementary Table S2.2.

Several pathways derived from different trait GWAS were recurrently enriched (Supplementary Table S3). For example, the serotonin uptake pathway was associated with iRBD when using PRS derived from AD, Bipolar disease (BD), and LBD GWAS. Major depressive disorder (MDD) and PTSD GWAS–derived pathways shared multiple enriched pathways, including the MHC pathway, antigen processing and presentation of exogenous peptide antigen via MHC class II, cell adhesion molecules, and MHC class II antigen presentation pathways.

### Burden Analysis provides further support for the association of multiple pathways

To investigate the contribution of rare variants within the 58 pathways associated with RBD risk identified in the previous step, we performed a burden test. Among the pathways analyzed, 20 showed nominal associations with RBD (p < 0.05). None remained significant after correction for multiple testing (Table 1, Supplementary Table S4).

**Table 1.** Nominally significant rare variant associations in pathway-level burden analysis (p < 0.05). No pathways remained significant after multiple testing correction. The complete list of results provided in Supplementary Table S5.

| Pathway | TestT<br>ype | SetID | Pvalu<br>e | FDR |
| --- | --- | --- | --- | --- |
| GOBP_SEROTONIN_TRANSPORT | skato | LoF | 1.57E-03 | 0.26 |
| GOBP_SEROTONIN_TRANSPORT | skato | functional | 1.89E-03 | 0.26 |
| GOBP_CHAPERONE_MEDIATED_AUTOPHAGY | burden | LoF | 1.95E-03 | 0.26 |
| HP_NEUROINFLAMMATION | burden | rare | 2.35E-03 | 0.26 |
| LYSOSOME | skato | LoF | 2.47E-03 | 0.26 |
| BIOCARTA_GABA_PATHWAY | burden | rare | 3.79E-03 | 0.30 |
| HP_NEUROINFLAMMATION | skato | rare | 3.95E-03 | 0.30 |
| GOBP_CHAPERONE_MEDIATED_AUTOPHAGY | skato | LoF | 4.47E-03 | 0.30 |
| BIOCARTA_GABA_PATHWAY | skato | rare | 5.63E-03 | 0.33 |
| HP_ABNORMAL_SUBSTANTIA_NIGRA_MORPHOLOGY | skato | functional | 9.25E-03 | 0.50 |
| GOBP_CHAPERONE_MEDIATED_AUTOPHAGY | burden | functional | 1.09E-02 | 0.53 |
| GOMF_MHC_CLASS_II_RECEPTOR_ACTIVITY | skato | LoF | 1.35E-02 | 0.58 |
| REACTOME_MHC_CLASS_II_ANTIGEN_PRESENTATION | burden | rare | 1.54E-02 | 0.58 |
| BIOCARTA_AGPCR_PATHWAY | burden | LoF | 1.87E-02 | 0.58 |
| GOBP_AUTOPHAGOSOME_LYSOSOME_FUSION | burden | rare | 1.99E-02 | 0.58 |
| GOBP_NEGATIVE_REGULATION_OF_IMMUNE_RESPONSE | skato | NS | 2.04E-02 | 0.58 |
| GOCC_GABA_RECEPTOR_COMPLEX | skato | CADD | 2.09E-02 | 0.58 |
| GOBP_CHAPERONE_MEDIATED_AUTOPHAGY | skato | functional | 2.27E-02 | 0.58 |
| GOBP_NEGATIVE_REGULATION_OF_AUTOPHAGY | burden | CADD | 2.53E-02 | 0.58 |
| GOBP_AUTOPHAGOSOME_LYSOSOME_FUSION | skato | rare | 2.56E-02 | 0.58 |
| BIOCARTA_AGPCR_PATHWAY | skato | LoF | 2.66E-02 | 0.58 |
| GOMF_ANTIGEN_BINDING | skato | LoF | 3.12E-02 | 0.58 |
| BIOCARTA_GABA_PATHWAY | burden | CADD | 3.17E-02 | 0.58 |
| GOBP_SEROTONIN_TRANSPORT | skato | NS | 3.32E-02 | 0.58 |
| GOBP_ANTIGEN_PROCESSING_AND_PRESENTATION_OF_PEPTIDE_OR_POLYSACCHARIDE_ANTIGEN_VIA_MHC_CLASS_II | burden | NS | 3.45E-02 | 0.58 |
| GOBP_DOPAMINE_SECRETION | burden | NS | 3.54E-02 | 0.58 |
| LYSOSOME | burden | rare | 3.60E-02 | 0.58 |
| REACTOME_ACETYLCHOLINE_NEUROTRANSMITTER_RELEASE_CYCLE | skato | NS | 3.97E-02 | 0.58 |
| WP_GABA_RECEPTOR_SIGNALING | burden | rare | 4.15E-02 | 0.58 |
| BIOCARTA_BLYMPHOCYTE_PATHWAY | skato | rare | 4.47E-02 | 0.58 |
| WP_GABA_RECEPTOR_SIGNALING | skato | CADD | 4.55E-02 | 0.58 |
| GOBP_POSITIVE_REGULATION_OF_CALCIUM_ION_TRANSPORT | burden | NS | 4.57E-02 | 0.58 |
| BIOCARTA_BLYMPHOCYTE_PATHWAY | burden | rare | 4.60E-02 | 0.58 |
| HP_ABNORMAL_SUBSTANTIA_NIGRA_MORPHOLOGY | skato | LoF | 4.62E-02 | 0.58 |
| GOBP_SEROTONIN_TRANSPORT | skato | CADD | 4.71E-02 | 0.58 |
| GOMF_ANTIGEN_BINDING | burden | LoF | 4.77E-02 | 0.58 |

**FDR**, false discovery rate; **LoF**, loss-of-function; **CADD**, Combined Annotation Dependent Depletion score; **NS**, nonsynonymous.

### Leave-one-gene-out and Gene-based burden analysis identify novel genes potentially associated with iRBD

To explore potential key drivers of the nominal associations observed in the pathway-level burden analysis, leave-one-gene-out analyses were performed. Overall, 117 genes were identified as potentially driving the observed pathway-level signals (Supplementary Table S5). Subsequently, we conducted a gene-based burden analysis to directly assess the contribution of these individual genes to iRBD risk, allowing us to nominate candidate genes suggested by the pathway-level analyses.

The gene-based burden analysis of these 117 candidate genes revealed 20 genes with nominal associations (p < 0.05), of which five genes, *GBA1*, *PLEKHM1*, *LRP2*, *P2RX1*, and *HAP1*, remained significant after FDR correction (Table 2). These results demonstrate robust gene-level associations and support the contribution of these genes to the observed pathway-level signals.

**Table 2.** Twenty genes showed nominal significance, with five genes passing FDR significance in the gene-based burden analysis. The complete list of gene-based burden results is provided in Supplementary Table S6.

| Gene | Method | SetID | N.Marker.Test | P.value | FDR |
| --- | --- | --- | --- | --- | --- |
| PLEKHM1 | skato | rare | 1136 | 6.46E-07 | **5.23E-05** |
| PLEKHM1 | burden | rare | 1136 | 7.81E-07 | **5.23E-05** |
| GBA1 | skato | LoF | 6 | 9.26E-04 | **3.74E-02** |
| LRP2 | skato | LoF | 52 | 1.27E-03 | **3.74E-02** |
| P2RX1 | skato | LoF | 15 | 1.64E-03 | **3.74E-02** |
| HAP1 | burden | NS | 42 | 1.67E-03 | **3.74E-02** |
| ITGB3 | skato | CADD | 25 | 4.14E-03 | 7.72E-02 |
| CACNB3 | burden | NS | 20 | 5.07E-03 | 7.72E-02 |
| TREM2 | skato | NS | 13 | 5.66E-03 | 7.72E-02 |
| ITGB3 | skato | NS | 36 | 5.76E-03 | 7.72E-02 |
| PRKAR1B | skato | LoF | 6 | 1.50E-02 | 1.83E-01 |
| NOS1 | skato | CADD | 32 | 2.08E-02 | 2.30E-01 |
| SNCAIP | skato | LoF | 7 | 2.23E-02 | 2.30E-01 |
| LILRA2 | burden | LoF | 8 | 2.45E-02 | 2.35E-01 |
| GABRA6 | burden | CADD | 11 | 2.93E-02 | 2.44E-01 |
| LILRB1 | skato | NS | 54 | 3.08E-02 | 2.44E-01 |
| HLA-DMA | burden | NS | 17 | 3.35E-02 | 2.44E-01 |
| LILRA2 | skato | LoF | 8 | 3.42E-02 | 2.44E-01 |
| GABBR1 | burden | NS | 22 | 3.53E-02 | 2.44E-01 |
| GABBR2 | skato | CADD | 32 | 3.94E-02 | 2.44E-01 |
| NOS1 | skato | NS | 47 | 3.98E-02 | 2.44E-01 |
| PRKAR1B | burden | LoF | 6 | 4.14E-02 | 2.44E-01 |
| BMF | burden | CADD | 22 | 4.26E-02 | 2.44E-01 |
| GRK4 | skato | LoF | 13 | 4.90E-02 | 2.44E-01 |
| SYT1 | skato | NS | 9 | 4.96E-02 | 2.44E-01 |

***N.Marker.Test***, number of marker tested; ***FDR***, false discovery rate; ***LoF***, loss-of-function; ***CADD***, Combined Annotation Dependent Depletion score; ***NS***, nonsynonymous.

Notably, LoF and functional variant burdens in the serotonin transport pathway, as well as LoF burdens in the chaperone-mediated autophagy (CMA) pathway, were among the top nominally associated pathways in the burden analysis. These pathways also ranked among the strongest signals in the pathway-specific PRS analysis, passing stringent FDR correction in both analytic frameworks. Importantly, the associations observed for functional variants in the serotonin transport pathway and LoF variants in the CMA pathway were not driven by a single dominant gene, indicating broader, polygenic contributions across multiple genes within each pathway. Within the serotonin transport pathway (LoF variant set), *P2RX1* emerged as a key contributor, with its LoF variants demonstrating significant association in the gene-based burden tests.

Since the role of *GBA1* is strongly established in PD and DLB, we further evaluated the association of rare variants in *PLEKHM1*, *LRP2*, *P2RX1*, and *HAP1* in PD and DLB cohorts (Supplementary Table S7 and S8). Of these four genes, *LRP2*, *P2RX1*, and *HAP1* showed nominal significance across different variant categories.

## Discussion

In this study, we systematically characterized the genetic architecture of iRBD by integrating pathway-specific PRS derived from neurodegenerative disorders and psychiatric traits and with rare variant burden analyses. Through this approach, we identified several pathways in which genetic risk originating from neurodegenerative and psychiatric disorders may contribute to iRBD susceptibility. In parallel, our rare variant analyses highlighted specific pathways, as well as individual genes that may influence iRBD risk. Together, this integrative strategy enhances our understanding of the genetic landscape of iRBD and provides insight into key biological processes and candidate genes that warrant further functional investigation.

The strong enrichment of pathways related to neurotransmission and synaptic function, derived from neurodegenerative disorder GWAS, aligns with the well-established role of these neurotransmitter systems in the pathophysiology of neurodegenerative diseases^12^. Among the disorders analyzed, pathways derived from LBD GWAS showed the greatest enrichment, consistent with our genetic correlation results indicating that iRBD is potentially more closely related to LBD than to AD or PD, supporting previous findings^13^.

Among psychiatric disorders, PTSD demonstrated substantial enrichment, with many pathways involved in immune response. This is consistent with a previous study, showing that general PTSD PRS (i.e. not pathway specific) is associated with increased iRBD susceptibility^14^, and with previous studies suggesting an epidemiological link between PTSD and RBD^15, 16^. The effect of several pathways on iRBD was shared between PTSD and MDD, including the MHC pathway, antigen processing and presentation via MHC class II, and cell adhesion molecules, highlighting immune dysregulation as a potential contributor to iRBD risk. Other psychiatric disorders, including anxiety, BD, insomnia, and problematic alcohol use (PAU), showed heterogeneous patterns, reflecting distinct molecular mechanisms underlying their potential comorbidity with RBD. These findings provide genetic evidence for the clinical observation of increased RBD comorbidity in psychiatric populations^8, 9, 10^.

Interestingly, several pathways were recurrently enriched across both neurodegenerative and psychiatric disorders. For example, serotonin uptake pathways derived from AD, BD, and LBD GWAS were associated with iRBD, suggesting convergent molecular mechanisms that may predispose individuals to RBD and neurodegeneration. The convergence of evidence from both rare variant and polygenic risk analyses specifically underscores the potential importance of serotonin transport in iRBD pathogenesis. Reduced motor neuron excitation can reinforce REM sleep atonia, and loss of serotonergic activity during REM sleep may also function to reduce motor neuron activity excitation and thereby reinforce REM atonia^17^. Thus, it is possible that motor behaviors in iRBD could arise, at least in part, from abnormal dysregulation of serotonin transport. Another pathway supported by association from multiple disorders is CMA, whose dysfunction has been shown to broadly contribute to the accumulation of toxic proteins in multiple neurodegenerative diseases, such as α-synuclein in PD^18^, amyloid-β and tau in AD^19, 20^, and TDP-43 (TAR DNA-binding protein 43) in ALS^21^. Together, these findings suggest that both serotonin dysregulation and impaired protein clearance may contribute to RBD pathogenesis.

Gene-level burden analyses identified five genes in which rare variants were robustly associated with RBD: *GBA1*, *PLEKHM1*, *LRP2*, *P2RX1*, and *HAP1*. *GBA1* (glucosylceramidase beta 1) encodes the lysosomal enzyme glucocerebrosidase, which plays a critical role in lipid degradation and lysosomal homeostasis^22^. Variants in *GBA1* have been identified in RBD genome-wide association studies (GWAS) ^13^, and in full sequencing of *GBA1*^23^, reinforcing its established link to iRBD. Other genes also have plausible mechanistic connections to the disorder. *PLEKHM1* (pleckstrin homology domain-containing family M member 1) is highly expressed in brain and whole blood based on GTEx data (Supplementary Fig. 1) and plays a key role in lysosome fusion^24^. It has been linked to PD^25^, and mouse models show that overexpression of PLEKHM1 impairs autophagy, subsequently promoting αSyn-induced neurodegeneration and neuroinflammation^26^. These findings suggest that impaired lysosomal dynamics may contribute to RBD pathogenesis. *LRP2* (LDL receptor related protein 2) is highly expressed in brain (Supplementary Fig. 2). High levels of Lrp2 have been implicated to serve neuroprotective roles in AD^27^ and repressed lrp2 translation in mouse model leads to cell apoptosis in AD^28^, indicating that LRP2 may play a role in maintaining neuronal integrity. *HAP1* (huntingtin-associated protein 1), expressed in the brain (Supplementary Fig. 3), is involved in vesicular trafficking and endocytosis of membrane-bound cargo^29, 30^, processes that are critical for neuronal function and survival^31^ and have been implicated in PD^32, 33, 34^. Collectively, these findings suggest that multiple biological pathways, including lysosomal function, cholesterol and lipid metabolism, synaptic trafficking, and neuronal signaling, may contribute to iRBD pathogenesis. Notably, several of these genes also showed nominally significant associations in PD and DLB, suggesting partial overlap in genetic risk across these synucleinopathies.

Although nonsynonymous variants in *TREM2* (P = 5.66 × 10⁻³, FDR-adjusted P = 0.08) and LoF variants in *SNCAIP* (P = 2.23 × 10⁻^2^, FDR-adjusted P = 0.23) did not survive FDR correction and only showed nominally significant associations with, they warrant further attention in RBD. *TREM2* (Triggering receptor expressed on myeloid cells 2) is a membrane glycoprotein that plays a crucial role in regulating immune responses, phagocytosis, lipid metabolism, and inflammation^35, 36^. *TREM2* has been verified as a risk factor for AD, particularly late-onset forms^37^, and possibly in PD^36^. *SNCAIP* encodes Synphilin-1 which has been linked to PD. Synphilin-1 is a major α-synuclein–interacting protein and major component of the Lewy bodies found in neurons of PD patients^38^. Synphilin-1 has protective effects in mouse models of α-synuclein–related disease^39, 40^. Accordingly, LoF variants in *SNCAIP* may similarly impair synphilin-1–mediated protective pathways, potentially leading to early α-synuclein dysregulation. This highlights a plausible mechanistic link between *SNCAIP* variants and iRBD.

Although our study benefits from large, well-characterized cohorts and complementary analytic approaches, several limitations should be noted. First, our findings lack an independent replication cohort, which limits the ability to confirm the robustness and generalizability of the observed associations. Moreover, because the burden analysis did not identify any pathways that survived multiple testing correction, some of the nominally significant signals may reflect cohort-specific effects, technical variation, sampling bias or chance, rather than true biological associations. Consequently, these results should be interpreted with caution, and validation in independent populations will be necessary to establish the reproducibility and biological relevance of the observed trends. Second, our analyses were restricted to individuals of European ancestry, limiting generalizability to other populations. Third, functional validation of the identified pathways and genes is necessary to confirm their potential roles in iRBD pathophysiology. Future studies integrating transcriptomic, proteomic, and single-cell data may provide additional mechanistic insights and facilitate the development of targeted interventions.

In conclusion, our findings extend the current understanding of genetics and pathogenesis of iRBD and provide novel insights into the genetic connections between RBD and other neurologic and psychiatric traits. Furthermore, the identification of specific genes harboring rare variants offers potential targets for future functional studies.

## Methods

### Study population

Pathway-specific PRS analyses were performed in 1,573 iRBD cases and 16,022 controls after quality control (QC). Cases were recruited by the International RBD Study Group and genotyped at McGill University. Controls were drawn from the International RBD Study Group and the UK Biobank (UKB), with UKB controls restricted to individuals without neurodegenerative (codes G00-G99) or psychiatric disorders (codes F00-F99) based on International Classification of Diseases version-10 (ICD-10, field 41270) and matched to cases by age and sex. All participants were unrelated individuals of European ancestry. iRBD cases had a mean age of 58.6 ± 12.7 years and were 81.7% male; controls had a mean age of 57.3 ± 9.1 years and were 77.5% male.

Burden analyses were conducted in 1,264 iRBD cases and 2,581 controls after QC. iRBD cases were collected through international collaborators and sequenced at McGill University, while controls were obtained from the Accelerating Medicines Partnership – Parkinson’s Disease (AMP-PD) consortium and sequenced at the National Institutes of Health by the Center for Alzheimer’s and Related Dementias (CARD) laboratory. iRBD cases had a mean age of 66.9 ± 9.2 years and were 82.1% male; controls had a mean age of 69.9 ± 13.4 years and were 50.7% male.

iRBD, defined as RBD diagnosed prior to the onset of overt neurodegeneration, was established according to the International Classification of Sleep Disorders (2nd or 3rd Edition) criteria with video polysomnography^41^. All participants provided written informed consent, and study protocols were approved by the institutional review boards.

### Sample Quality Controls

For the pathway PRS analysis, McGill cohort samples were filtered prior to imputation based on heterozygosity (−0.15 ≤ F ≤ 0.15), call rate (>95%), and sex concordance, and related individuals closer than third-degree (pi-hat > 0.125) were excluded. Population structure was assessed using principal component analysis (PCA), and only individuals of European ancestry were retained. For the UKB controls, imputed genotyping data were obtained from unrelated individuals of European ancestry (field 22006) and randomly selected to match cases in age and gender, after excluding participants diagnosed with psychiatric disorders (ICD-10 codes F00–F99) or nervous system diseases (ICD-10 codes G00–G99; field 41270), with European ancestry confirmed via PCA.

For the burden analyses, samples were filtered based on genotype quality (GQ ≥ 20), depth of coverage (DP ≥ 10), and allele balance or variant allele fraction thresholds for heterozygous and homozygous calls according to GATK best practices; samples with excess heterozygosity (−0.15 ≤ F ≤ 0.15), >5% missing genotypes, or discordant genetic and reported sex were removed.

### Genotype Callset and Data Cleaning

Genotyping for the McGill cohort was conducted using the OmniExpress or NeuroBooster arrays, following the manufacturer’s protocols (Illumina Inc.). Prior to imputation, SNPs were filtered for missingness (<5%), differential missingness between cases and controls (p > 1 × 10⁻⁴), haplotype-based missingness (p > 1 × 10⁻⁴), and deviation from Hardy–Weinberg equilibrium (HWE) in controls (p > 1 × 10⁻⁴). Genotype imputation was carried out on the TOPMed Imputation Server using the TOPMed r3 reference panel with default parameters. The UK Biobank genetic dataset comprises 487,409 samples, which were phased and imputed using the Haplotype Reference Consortium and the combined UK10K + 1000 Genomes reference panels ^42^. For merging the McGill and UKB datasets, high-quality imputed variants (R² > 0.8) common to both cohorts were retained. Variants from each cohort were independently filtered based on minor allele frequency (MAF > 0.01), genotype missingness (GENO < 0.01), and HWE in controls (p > 5 × 10⁻⁶). After merging variants common to both cohorts, the same quality control filters were reapplied to ensure dataset integrity.

For whole-genome sequencing data, variants were filtered for GQ ≥ 20, DP ≥ 10, and allele number ≥ 95%. Variants deviating from HWE in controls (p < 1 × 10⁻⁶) or with >5% missing genotypes were excluded. Additional filters removed variants with high haplotype-level missingness (threshold = 0.0001). Joint variant calling was performed using DeepVariant–GLnexus. Only rare variants with a MAF < 0.01 were retained.

### Pathway Definition

Pathways were selected using a literature-informed approach, focusing on biological processes implicated in the pathophysiology of RBD (full list provided in Supplementary Table 9). A total of 279 pathways across 10 broad biological categories were included: (1) Neurotransmission and Synaptic Function, (2) Autophagy and Lysosomal Function, (3) Apoptosis and Cell Death, (4) Calcium Signaling, (5) Genetic and Antigen Presentation, (6) Neurodevelopment and Differentiation, (7) Immune Response and Inflammation, (8) Circadian Rhythm and Sleep, (9) Neurodegenerative and Neuropsychiatric Disease Related, and (10) Mitochondrial Function and Oxidative Phosphorylation^1, 13, 43, 44, 45, 46, 47, 48, 49, 50, 51, 52, 53, 54, 55^.

All pathway collections available in the Molecular Signatures Database (MSigDB) (https://www.gsea-msigdb.org/gsea/msigdb) were queried using relevant keywords (e.g., “neuroinflammation”) to identify biologically meaningful pathways potentially implicated in RBD. Pathways containing fewer than 10 genes were not included to ensure sufficient coverage for polygenic risk score (PRS) calculation. This filtering step minimized the risk of underpowered analyses due to sparse gene sets.

### PRS Analysis

Pathway-specific PRS were computed using PRSice-2^56^ and were constructed using the most recently released GWAS summary statistics from multiple diseases, including five neurodegenerative diseases: AD^57^, ALS^58^, LBD^59^, MSA^60^, and PD^61^—and seven psychiatric traits: anxiety^62, 63^, BD^63^, insomnia^64^, MDD^65^, PAU^66^, PTSD^67^ and SCZ^68^.

Standard QC was performed on the GWAS summary statistics before PRS construction. Briefly, SNPs with a minor allele frequency (MAF) < 1% and imputation INFO score < 0.8 (when available) were excluded. Linkage disequilibrium (LD) clumping was applied using an LD threshold of r² > 0.1 within a 1 Mb window. The regression models were adjusted for age at onset (or age at diagnosis when onset age was unavailable) for cases, age at enrollment for controls, sex, and the top two PCs, selected based on a Scree plot (Supplementary Fig. 4), as covariates. We performed 10,000 permutations to generate empirical p-values for evaluating pathway enrichment. Pathways with fewer than four SNPs contributing to PRS construction were excluded prior to FDR correction.

### Rare Variant Burden Analysis

The optimized Sequence Kernel Association Test (SKAT-O, R package)^69^ was used to analyze the joint effects of rare variants. SKAT-O evaluates the aggregated effects of rare variants within predefined genomic sets, accounting for both the direction and magnitude of individual variant effects. Analyses were restricted to pathways that reached nominal significance (p < 0.05) in the previous pathway-specific PRS analysis. Gene coordinates were defined using MANE Select transcripts, with an additional 300 base pairs upstream and downstream. Variants were annotated using the Variant Effect Predictor (VEP)^70^. Burden analyses were conducted across five variant categories: (1) all rare variants, (2) functional variants, defined as nonsynonymous, splice-site, frameshift, and stop-gain variants, (3) loss-of-function variants, including splice-site, frameshift, and stop-gain variants, (4) nonsynonymous variants, and (5) predicted deleterious variants, defined as variants with a Combined Annotation Dependent Depletion (CADD) score >20, representing the top 1% of predicted deleterious variants. The regression model was adjusted for age, sex, and the first five principal components, selected based on a Scree plot (Supplementary Fig. 5), to account for population stratification.

To assess whether pathway-level signals were driven by individual genes, a leave-one-gene-out analysis was performed in which each gene was sequentially removed and the burden test repeated. A pathway was considered gene-driven if the p-value shifted from significant to non-significant after removal of a single gene. Genes identified as potential drivers were subsequently evaluated using gene-based burden analysis for confirmation. To ensure robust results, genes comprising fewer than four SNPs in the burden analysis were excluded before applying FDR correction.

### Rare Variant Burden Analysis in PD and DLB

Whole-genome sequencing data for PD and DLB were obtained from the AMP-PD Study Group (v4 release; https://amp-pd.org/). Initial quality control was performed by AMP-PD as previously described (https://amp-pd.org/whole-genome-data). Additional quality control was applied, including filtering variants with depth of coverage (DP) ≤ 25, removing samples with >5% missing genotypes, excluding individuals with excess heterozygosity (F < −0.15 or F > 0.15), and removing samples with call rate <95%. Individuals of non-European ancestry and those related closer than third-degree (pi-hat > 0.125) were also excluded. After filtering, the dataset included 3,051 PD cases, 3,667 PD controls, 2,605 DLB cases, and 1,894 DLB controls. An additional PD cohort from the UKB was included, for which the same quality control procedures were applied, except that both GQ ≥ 25 and DP ≥ 25 thresholds were enforced. This cohort consisted of 3,173 PD cases and 31,722 controls without proxy cases, and 18,110 PD cases and 180,961 controls when proxy cases were included.

Rare variant burden analyses in PD and DLB were restricted to genes identified as significantly associated with iRBD in the gene-based rare variant analyses and were conducted using the same statistical framework and covariate adjustments as described above. Meta-analysis across the PD cohorts from AMP-PD and UK Biobank was performed using MetaSKAT ^71^.

## Data Availability

AD, ALS, LBD and MSA summary statistics are publicly available on the NHGRI-EBI GWAS Catalog (https://www.ebi.ac.uk/gwas/) under accession numbers GCST90027158, GCST90027164, GCST90001390, and GCST90406924, respectively. PD summary statistics are available from the NDPK portal (https://ndkp.hugeamp.org/research.html?pageid=a2f_downloads_280). All psychiatric disorder GWAS summary statistics are available via the Psychiatric Genomics Consortium (PGC) website (https://pgc.unc.edu/for-researchers/download-results/), except insomnia (https://vu.data.surfsara.nl/index.php/s/06RsHECyWqlBRwq) and PAU (https://yale.app.box.com/s/m4tg6s87zor1bp3uldkrro2xnezg6kul)

## Code Availability

The code for all analyses is publicly available on https://github.com/MerZ03/RBD_neuro_psy.

## Acknowledgements

We would like to thank the research participants for contributing to this study. This study was financially supported through grants from the Hilary and Galen Weston Foundation, Michael J. Fox Foundation (MJFF) and the Canadian Consortium on Neurodegeneration in Aging (CCNA). Additionally, the G-Can (GBA1-Canada) Initiative, an open-science collaborative initiative aimed at addressing *GBA1*-associated neurodegeneration, has made contributions to this research. G-Can is supported by The Hilary and Galen Weston Foundation, Silverstein Foundation, and J. Sebastian van Berkom and Ghislaine Saucier. This work was supported in part by the Intramural Research Program of the National Institute on Aging (NIA), and the Center for Alzheimer’s and Related Dementias (CARD), within the Intramural Research Program of the National Institute on Aging and the National Institute of Neurological Disorders and Stroke (1ZIAAG000546) as well as NIH NIA grants R34 AG056639, U19 AG071754, P50 AG016574, P30 AG062677 and VA RRD RX004822. MS and KK received funding from the Slovak Research and Development Agency under contract no. APVV-22-0279, and the EU Renewal and Resilience Plan “Large projects for excellent researchers” under grant No. 09I03-03-V03-00007. The iRBD cohort study at First Medical Faculty (Prague, Czechia) is supported by the Czech Health Research Council (grants NU21-04-00535 and NW25-04-0079) and the National Institute for Neurological Research (Project No. LX22NPO5107), funded by the European Union – Next Generation EU. This research used the NeuroHub infrastructure and was undertaken thanks in part to funding from the Canada First Research Excellence Fund, awarded through the Healthy Brains, Healthy Lives initiative at McGill University, Calcul Québec and Compute Canada. This research has been conducted using the UK Biobank Resource under Application Number 45551. The UKB cohort was accessed using Neurohub (https://www.mcgill.ca/hbhl/neurohub). ZG-O is supported by the Fonds de recherche du Québec – Santé (FRQS) Chercheurs-boursiers award and is a William Dawson Scholar. Samples from the National Centralized Repository for Alzheimer’s Disease and Related Dementias (NCRAD), which receives government support under a cooperative agreement grant (U24 AG021886) awarded by the National Institute on Aging (NIA), were used in this study. We thank contributors who collected samples used in this study, as well as patients and their families, whose help and participation made this work possible. This research was funded in part by the Neuro Genomics Partnership (NGP). The NGP is a consortium supported by Takeda and Roche, who contributed to the funding of this study.

## Author Contributions

Z.G.-O. conceptualized and supervised the study, acquired funding, and contributed to review and editing. Z.Z. contributed to conceptualization, performed data analysis and interpretation, prepared the figures, and wrote and revised the manuscript. E.S. and Z.-H.F. performed RBD WGS data preparation and quality control. L.L. performed genotyping data preparation and quality control. F.A., J.A., S.A., and M.T. performed sample preparations and validation. P.D., I.A., M.T.M.H., A.D., Y.D., M.A.-S., A.I., A.S., B.H., C.G., A.M., G.M., A.I., M.S., G.L.G., M.V., F.J., A.B., K.S., D.K., P.D., M.S., S.R., M.F., M.P., B.M., C.T., F.S.-D., G.P., F.B., E.A., V.C.D.C., M.T., G.F., A.H., L.F.-S., M.O., M.S., K.K., J.B., B.A., B.O., P.M., D.A. contributed to sample collection. B.F.B., Y.-E.J., O.A.R., S.-Y.W., J.L., D.P., C.B., H.L., R.B.P., G.A.R. and Z.G.-O. supervised the study. All authors critically reviewed and edited the article.

## Corresponding Author

Correspondence to Ziv Gan-Or.

