## Supplemental Figures for "Shared Pathogenic Pathways Between REM Sleep Behavior Disorder and Neurodegenerative and Psychiatric Disorders"

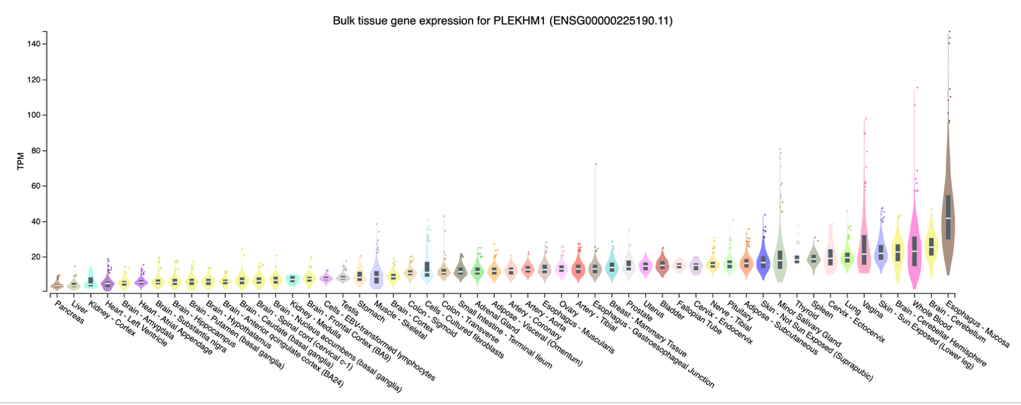

**Figure S1.** Gene expression data for *PLEKHM1* retrieved from the GTEx Portal (<https://gtexportal.org>, accessed Oct 16,2025)

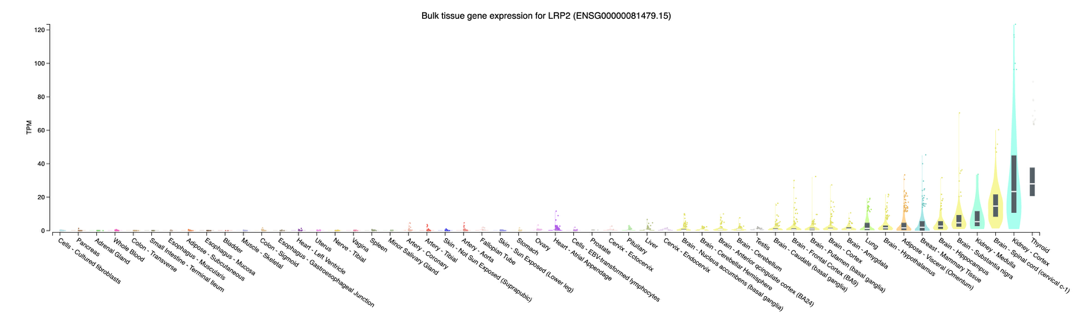

**Figure S2.** Gene expression data for *LRP2* retrieved from the GTEx Portal (<https://gtexportal.org>, accessed Oct 16,2025)

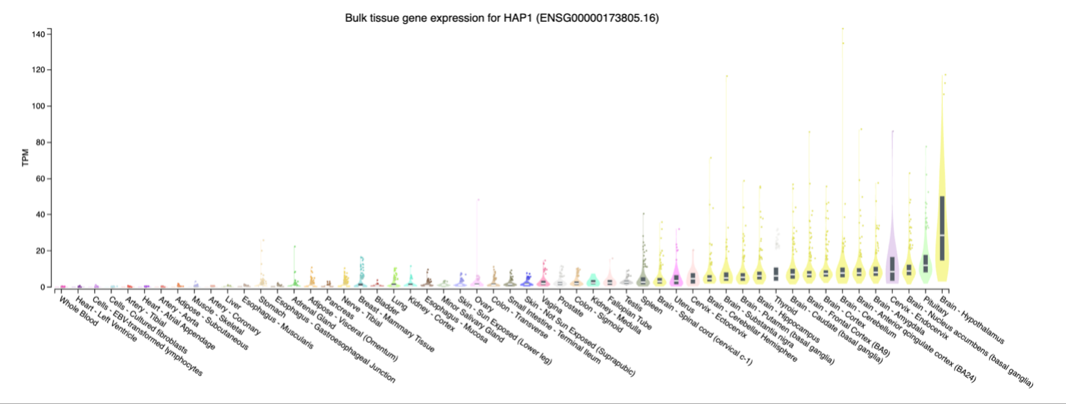

**Figure S3.** Gene expression data for *HAP1* retrieved from the GTEx Portal (<https://gtexportal.org>, accessed Oct 16,2025)

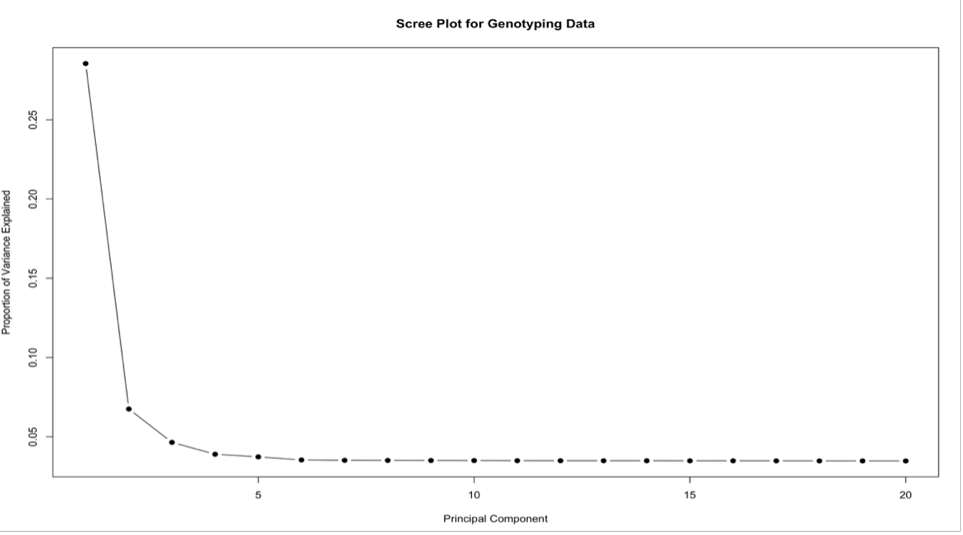

**Figure S4.** Scree plot from principal component analysis showing that the first two principal components capture the majority of variance in the genotyping data.

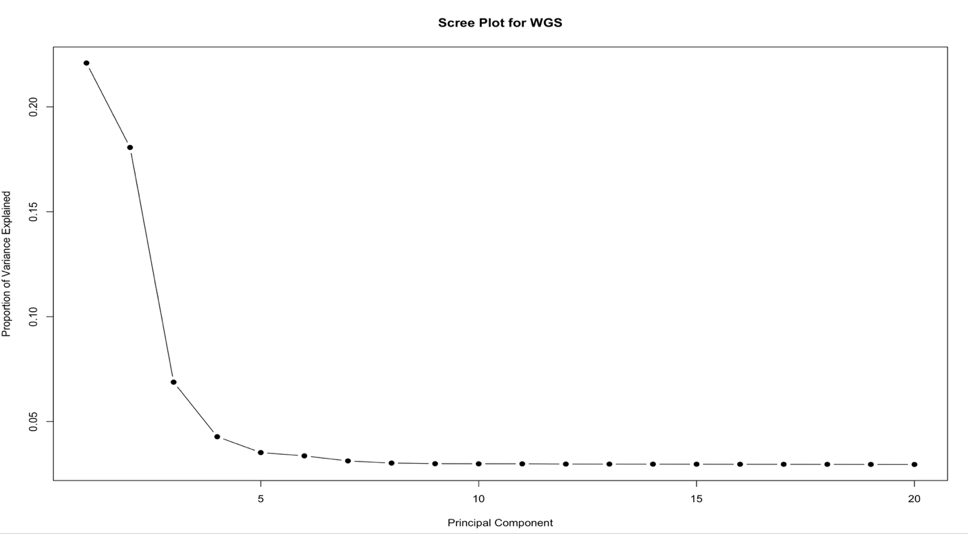

**Figure S5.** Scree plot from principal component analysis showing that the first two principal components capture the majority of variance in the WGS data.
